# A Simultaneous [^11^C]Raclopride Positron Emission Tomography and Functional Magnetic Resonance Imaging Investigation of Striatal Dopamine Binding in Autism

**DOI:** 10.1101/2020.08.10.20172197

**Authors:** Nicole R. Zürcher, Erin C. Walsh, Rachel D. Phillips, Paul M. Cernasov, Chieh-En J. Tseng, Ayarah Dharanikota, Eric Smith, Zibo Li, Jessica L. Kinard, Joshua C. Bizzell, Rachel K. Greene, Daniel Dillon, Diego A. Pizzagalli, David Izquierdo-Garcia, David Lalush, Jacob M. Hooker, Gabriel S. Dichter

## Abstract

**Background:** The social motivation hypothesis of autism suggests that autism spectrum disorder (ASD) is characterized by impaired motivation to seek out social experience early in life that interferes with the development of social functioning. This framework posits that impaired mesolimbic dopamine (DA) function underlies compromised responses to social rewards in ASD. Although this hypothesis is supported by functional magnetic resonance imaging (fMRI) studies, no molecular imaging study has evaluated striatal dopamine functioning in response to rewards in ASD.

**Methods:** This study evaluated striatal dopaminergic functioning during incentive processing in ASD using simultaneous positron emission tomography (PET) and fMRI using the D2/D3 dopamine receptor antagonist [^11^C]raclopride. Using a bolus + infusion protocol, voxel-wise binding potential (BP_ND_) was compared between groups (Controls=12, ASD=10) in the striatum.

**Results:** Relative to controls, the ASD group demonstrated relatively decreased phasic DA release to incentives in the right and left putamen and left caudate. Striatal clusters showing significant between-group BP_ND_ differences were used as seeds in whole-brain fMRI general functional connectivity analyses. This revealed increased connectivity between the PET-derived right putamen seed and clusters in the precuneus and right insula in the ASD group. Within the ASD group, decreased phasic DA release in the left putamen was related to poorer theory-of-mind skills.

**Conclusions:** ASD was characterized by impaired striatal phasic DA release and abnormally increased functional connectivity, providing support for the social motivation hypothesis of autism. PET measures of dopamine receptor target occupancy may be suitable to evaluate novel ASD therapeutics targeting the striatal dopamine system.

## Introduction

The social motivation hypothesis of autism proposes that functional disruptions in brain circuits supporting social motivation constitute a primary deficit that contributes to social communication impairments [1]. In particular, this framework posits that social communication symptoms in autism spectrum disorder (ASD) reflect decreased motivation to engage in reciprocal social behaviors throughout development that results in fewer experiences with social rewards [2]. When children with ASD lack the motivation to participate in activities where social skills are typically forged, the resulting impoverished social environment compounds social impairments and negatively impacts the development of social communication [3].

Social motivation is supported by the same substrates that govern other motivated behaviors, namely ascending dopamine (DA) projections from the ventral tegmental area to the striatum and prefrontal cortex, forming a DA pathway sensitive to reward magnitude and probability [4-6]. This DA system mediates responses to social and nonsocial incentives [7, 8], and striatal DA transmission influences social behaviors [9]. Numerous functional magnetic resonance imaging (fMRI) studies have reported that ASD is characterized by decreased striatal responses to rewards [10-14, e.g., 15], highlighting striatal involvement in impaired social motivation in ASD. Additional findings indicate that striatal DA dysfunction is implicated in the etiology of ASD [16, 17]. First, there is evidence of impaired striatal functioning in ASD in the form of altered effort-based decision-making for rewards [18]. Second, polymorphisms of the DA D4 receptor gene and the DA transporter gene are related to challenging behaviors [19] and repetitive behaviors [20] in ASD, and there are links between polymorphisms of the DA-3-receptor gene and striatal volumes and repetitive behaviors in ASD [21]. Furthermore, oxytocin abnormalities in ASD [22, 23], reports of the therapeutic effects of intranasal oxytocin administration for treating core ASD symptoms [24, 25], and the effects of oxytocin on striatal responses to rewards in ASD [26] support an etiologically-relevant role for mesolimbic DA functioning in ASD. Of note, there are dense oxytocin projections within the mesolimbic DA system, including oxytocin neurons that project to the ventral tegmental area and nucleus accumbens [27], and oxytocin receptor activation plays an important role in the activation of reward pathways during prosocial behaviors [28-30]. Finally, a widely used ASD preclinical model, the valproic acid model, [31, 32] causes a cascade of neurobiological changes including excitatory/inhibitory neural imbalances linked to increased basal DA in the frontal cortex [33], hyperactive mesocortical DA in response to stress [34], and changes in locomotor behavior akin to that observed in striatal DA-depleted animals [35].

Despite converging evidence supporting the involvement of striatal DA impairments in the pathophysiology of ASD, no molecular imaging study has investigated striatal DA functioning in ASD. The goal of this study was to use simultaneous fMRI and positron emission tomography (PET) with the D2/D3 dopamine receptor antagonist [^11^C]raclopride to investigate striatal functioning during incentive processing in ASD. Neutral and rewarding incentives were presented during a behavioral fMRI task, and a bolus+infusion [^11^C]raclopride PET paradigm allowed measurement of both dopaminergic tone and phasic dopaminergic release in response to incentives. We hypothesized that the ASD group would be characterized by decreased striatal phasic DA release in response to incentives relative to a control group, indexed by the nondisplaceable binding potential (BP_ND_) of [^11^C]raclopride. We also hypothesized that, compared to controls, the ASD group would exhibit abnormal functional connectivity, assessed by fMRI, between striatal seed regions that showed reduced phasic DA release and their functional targets. Finally, exploratory analyses examined relations between striatal BP_ND_ and symptom severity in the ASD group.

## Methods and Materials

### Participants

Procedures were in accordance with the ethical standards of the UNC-Chapel Hill (UNC) institutional research board and with the 1964 Helsinki declaration and its later amendments or comparable ethical standards. The PET protocol was approved by the UNC Radioactive Drug Research Committee. Participants with ASD and typically developing controls were recruited via the UNC Autism Research Registry and a university email listserv, respectively. Groups were matched on age, gender, and IQ. Participants with ASD were capable of providing informed consent and did not require surrogate consent. Exclusion criteria for both groups included lack of fluent phrase speech, IQ<70, known sensory deficits (blindness and deafness), or medical conditions, history of neurological injury, intellectual disability, and MRI or PET contraindications. The control group had no lifetime psychiatric diagnosis, assessed by the Structured Clinical Interview for DSM-5 (SCID-5-RV) [36]. The ASD group had no current diagnosis of substance abuse or mood disorders and no lifetime psychiatric diagnosis except for ASD, assessed by the SCID-5-RV [36].

Potential control participants completed the Social Communication Questionnaire (≤ 15 cutoff) to rule out possible ASD symptoms [37] and a screener for intellectual functioning (the North American Adult Reading Test [NAART, 38]). To aid group matching, control participants with NAART estimated IQ scores greater than 120 were excluded. Eligible participants completed an in-person assessment that included the SCID-5, the Wechsler Abbreviated Scale of Intelligence [WASI; 39], the self-report Social Responsiveness Scale, Second Edition (SRS-2), a dimensional measure of ASD symptoms (Constantino et al., 2003), and the “Reading the Mind in the Eyes” Test, Revised Version (RMITE) [40], a measure of theory-of-mind. The ASD group also completed module 4 of the Autism Diagnostic Observation Schedule-2 (ADOS-2) [41] administered by a reliable assessor (JLK & RKG) to confirm ASD diagnoses. Eligible participants were then scheduled for the PET-MR scan. Participants received $20/hour for the assessment and between $160 to $200 (amount based on behavioral task performance, described below) for the scan.

Twenty-six individuals with ASD and 34 controls (ages 19-29 years) provided written informed consent. Of these 60 potential participants, 23 were ineligible after in-person evaluation (11 controls, 12 with ASD) and one declined further participation. Of the 36 participants who completed scanning, data from 22 were analyzable: 14 participants were not included in the final sample due to problems with PET injection or scanner (4), incomplete/missing due to technical difficulties (8), abnormally low and noisy PET counts (1), and excessive motion during the PET scan (1). The final sample with analyzable PET data included 10 participants with ASD (all male; all white; 1 Hispanic) and 12 controls (10 males; 8 white, 2 Black or African American, 1 Asian, 1 race not reported; 2 Hispanic). Groups did not differ in gender, race, or ethnicity distributions, Fisher’s exact test *p*’s>0.34. As depicted in Table 1, the ASD group differed from controls with respect to scores on the SRS-2 and RMITE, but not IQ, SES, or age.

**Table 1.** Participant characteristics

|  | ASD (*n* = 10) | Control (*n* = 12) | Test statistic | *p*-value |
| --- | --- | --- | --- | --- |
|  | *M* (*SD*) | *M* (*SD*) |  |  |
| ADOS-2 CSS | 8.3 (2.1) | --- | --- | --- |
| SRS-2 Total T score | 67.6 (12.2) | 46.67 (7.2) | *t*(20) = 5.01 | <0.001** |
| WASI-2 Full Scale IQ | 116.5 (11.9) | 120.67 (8.9) | *t*(20) = 0.94 | 0.36 |
| RMITE | 22.6 (3.50) | 27 (2.30) | *t*(20) = -3.54 | 0.002* |
| SES | 38.3 (14.4) | 39.67 (11.5) | *t*(20) = 0.25 | 0.81 |
| Age | 24.9 (3.6) | 25.67 (4.3) | *t*(20) = 0.45 | 0.66 |
| Sex | 10 ♂ | 10 ♂, 2 ♀ | $\chi^2 = 0.22$ | 0.48 |

*Note.* ASD = Autism spectrum disorder group; Control = Control group; M = mean; SD = standard deviation; ADOS-2 CSS = Autism Diagnostic Observation Schedule, 2<sup>nd</sup> edition Calibrated Severity Score [56]; SRS-2 = Social Responsiveness Scale, Second Edition [88]; WASI-2 = Wechsler Abbreviated Scale of Intelligence, 2<sup>nd</sup> edition [39]; RMITE = Reading the Mind in the Eyes Test, Revised Version [40]; SES = Socioeconomic status measured by the Hollingshead Four Factor Index of Socioeconomic Status [89].
\*: *p*<.005; \*\*: *p*<.0001

### Simultaneous PET-MR scanning

Participants completed a simultaneous PET-MR scan on a Siemens Biograph mMR scanner at the UNC Biomedical Research Imaging Center using a bolus+infusion protocol with a planned K_bol_ of 105min [42]. PET acquisition took place for 63min. Approximately 1min after the PET scan began, a bolus injection of [^11^C]raclopride was administered after which the infusion injection of [^11^C]raclopride was administered using a Medrad® Spectris Solaris® EP MR Injection System (radioactivity was limited to 15mCi in total over the bolus and infusion and mass dose did not exceed 10µg (with a specific radioactivity at the bolus time of injection > 0.4 Ci/µmol)). A 6min structural T1-based MR sequence was obtained (FOV=256 mm, 1×1×1 mm resolution, TR=2530ms, TE=1.69ms, flip angle=7 degrees) for anatomical localization, spatial normalization of imaging data, and generation of attenuation correction maps [43], in addition to localizer and attenuation correction scans. Then, two resting-state scans were obtained (echo planar imaging, FOV=212 mm, 3.312×3.312×3.3 mm resolution, TR=3000, TE=30ms, flip angle=90 degrees). Next, three task blocks were presented during which fMRI data were collected simultaneously to the ongoing PET acquisition. See Figure 1 for timing of data collection, data modelling, and participant behavior.

**Figure 1.**
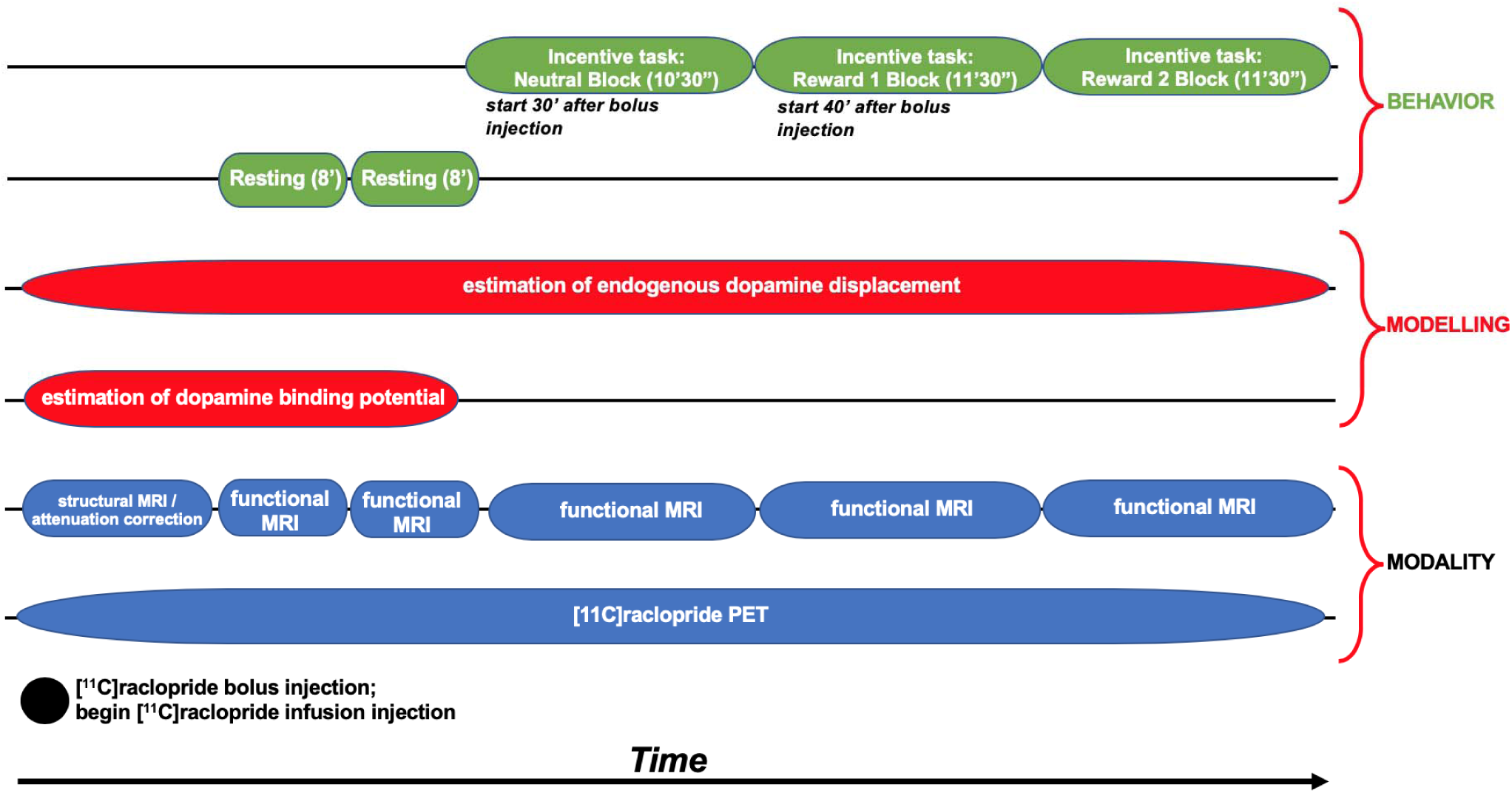
Timing of data collection, data modelling, and participant behavior during scanning.

### Behavioral Task During PET & fMRI scanning

Participants first completed two 8’09” resting-state fMRI runs with eyes open to allow for tracer uptake. Next, participants completed a monetary incentive delay task [44] modified for use in PET-MR studies. The task was presented using PsychoPy software version 1.84.1 [45]. The task presented a 10’30” neutral block followed by two 11’30” reward blocks. As shown in Figure 2, on each trial (6.37-15.17 sec), participants saw a blue polygon cue (1.5 sec), followed by a green circle target (0.367 sec) and an outcome (1.5 sec); these stimuli were separated by jittered inter-stimulus and inter-trial intervals during which a fixation cross was shown. The task required making a speeded button press with the right index finger upon seeing the target. During the neutral block, participants completed 63 trials that started with a square cue. No monetary rewards were delivered on these trials. Instead, sufficiently speeded button presses resulted in the presentation of a grey rectangle as a “no-reward” outcome. The other outcomes indicated either no response (“No Response!”), the response was too quick (within 100ms of the target presentation: “Too Fast!”), or it was made after an adaptive reaction time (RT) threshold (“Too Slow!”) that was programmed such that ~75% of each participant’s responses were successful.

The neutral block was followed by four reward runs, combined into two blocks (number of trials per reward run: block 1: 34/33, block 2: 33/34). The neutral and reward blocks were separated by a brief break. In the reward blocks, different polygon cues (square, triangle, pentagon, hexagon) indicated that trials could result in no-reward (grey rectangle) or a small (50 cents), medium (1 dollar), or large reward (5 dollars), respectively; the assignment of the four polygons to the four outcomes was stable across the reward blocks and counterbalanced across participants. Successful trials (i.e., trials with sufficiently speeded button presses) ended with images depicting the no-reward, small, medium, and large reward outcomes (Figure 2). Unsuccessful trials yielded the same feedback as in the neutral block (“Too Fast!”, “Too Slow!”, or “No Response!”). Following each neutral and reward block, participants rated cues and outcomes using a 9-point Likert scale with anchors of “very negative” and “very positive” at the ends and “neutral” in the center.

This MID task includes novel features designed to maximize detection of DA release in the PET-MR environment. First, the first reward block begins ~40min after the [^11^C]raclopride bolus injection after the target/reference ratio stabilized; the long uptake period serves as a baseline scan. Second, about 75% of reward trials result in reward feedback, including many $5 rewards; this success rate is higher and the large rewards are larger than in many MID studies to enhance incentive motivation, which should be evident in stronger signals related to reward anticipation and consummation [46]. Third, while most MID versions use explicit reward and neutral cues that make the potential outcome of each trial clear, the current design forces participants to learn which cues predict which reward magnitudes by experience. By adding associative learning, the current design should enhance sensitivity to positive prediction errors (and other learning-related signals) encoded by phasic DA release [6]. This modified version of the MID task was developed at McLean Hospital (by DD & DAP).

**Figure 2.**
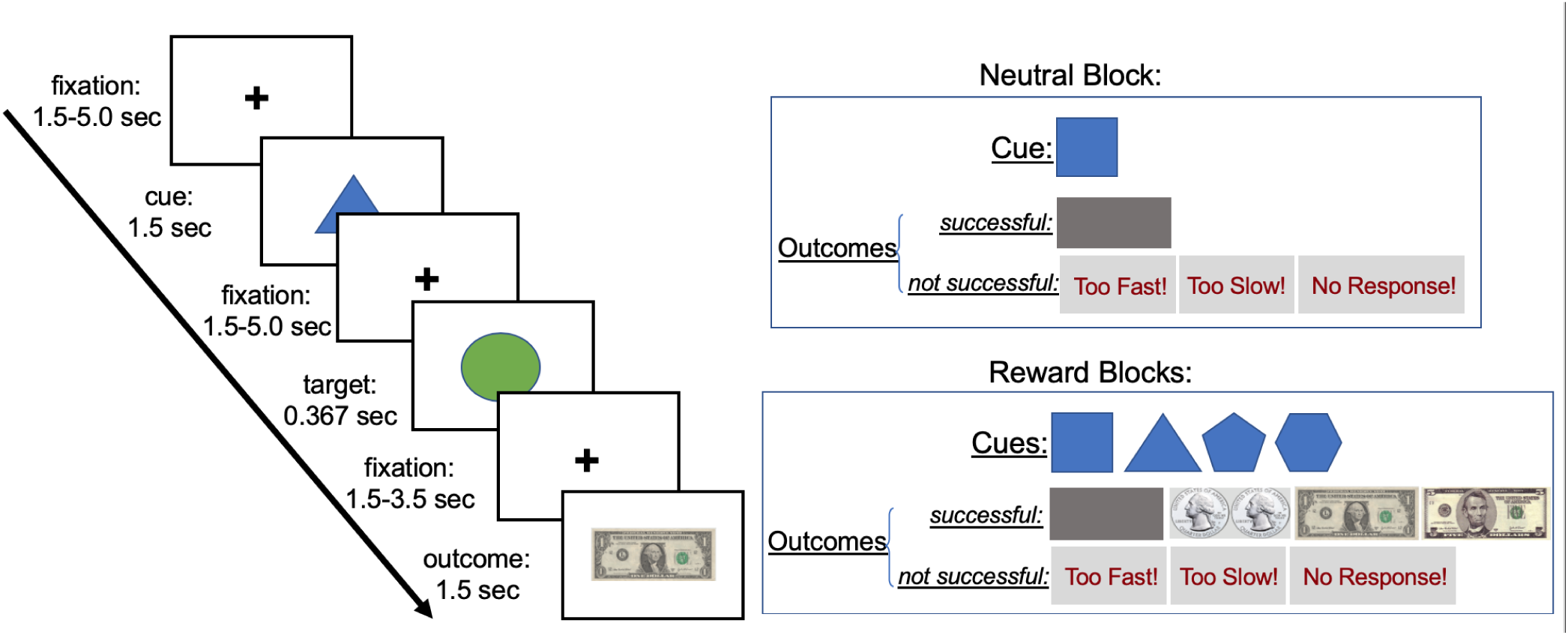
The PET-MR incentive task. Each trial consisted of a cue phase and an outcome phase. Trials were presented first in a neutral block that consisted of only neutral trials and then in two reward blocks that consisted of neutral trials and reward trials of varying magnitudes (small, medium, or large).

### PET Analysis

Post-scan reconstruction of the PET data used 1min frames [47]. BP_ND_ was defined as the ratio of selectively bound ligand to nondisplaceable ligand in the tissue at equilibrium using the two-part simplified reference tissue model (SRTM). A voxel-wise map of ASD>Control [^11^C]raclopride BP_ND_ (reward>neutral) values was created with a threshold of z-values>2.3 (i.e., *p*<0.012), which was subsequently masked by the bilateral caudate nucleus, putamen, and nucleus accumbens regions from the Harvard-Oxford probabilistic atlas (thresholded at 25% and binarized). Reduced BP_ND_ is interpreted to mean an increase in endogenous DA (i.e., competition with [^11^C]raclopride). However, reduced BP_ND_ could also represent decreased binding affinity due to receptor trafficking (i.e., reduced binding affinity due to receptor internalization). Results for the contrast of ASD>Control, reward>neutral, signify increased BP_ND_ or decreased phasic DA release to the reward condition, relative to the neutral condition, in the ASD group compared to controls. For a complete description of PET analyses see **Supplemental Materials I** and Sander and colleagues [48].

### fMRI General Functional Connectivity (GFC) Analysis

We used general functional connectivity (GFC) to examine whole-brain connectivity with striatal seed regions in which we observed significant differences in BP_ND_ between diagnostic groups. GFC, a method that combines resting-state and task fMRI data, offers better test-retest reliability and higher estimates of heritability than intrinsic connectivity estimates from the same amount of resting-state data alone [49]. In the present study, where combining the two resting-state runs and three task blocks yields 49’30” of fMRI data for connectivity analyses, GFC also offers the advantage of longer durations of fMRI data to be analyzed. This is critical given that more than 25min of fMRI data are needed to reliably detect individual differences in connectivity [50-52].

Voxel-wise whole-brain connectivity was evaluated using the CONN Toolbox’s seed-to-voxel analysis. Functional images were preprocessed with the default preprocessing pipeline in the SPM12 CONN functional connectivity toolbox, version 19c [53]. Steps included: resampling to 2×2×2-mm voxels and unwarping, centering, slice time correction, normalization to MNI template, outlier detection (ART-based scrubbing), and smoothing to an 8mm Gaussian kernel. Motion parameters were entered as multiple regressors and images with framewise displacement above 0.5mm or global BOLD signal changes above 3 SD were flagged as potential outliers and regressed out [54]. For the majority of the sample (n=19) all five runs of functional data were analyzable and all participants had at least 3 analyzable runs. Reasons for excluded runs: technical errors (2), striation artifacts (1), and excessive motion (2). There were no significant differences between groups on average motion, t(20)=0.35, *p*>.05, or average global BOLD signal changes, t(20)=1.07, *p*>.05.

### Supplementary fMRI Activation and Task-Based Generalized Psychophysiological Interactions (gPPI) Analyses

The main objectives of this study were to investigate striatal DA release in ASD and general functional connectivity with striatal regions showing impaired DA release in ASD. Therefore, fMRI activation methods are presented in **Supplemental Materials II** and secondary analyses using task-based gPPI are described in **Supplemental Materials III**.

## Results

### PET Results

[11C]raclopride dose did not differ between groups; for the ASD and control group, the mean (SD) dose was 12.39 (0.98) mCi and 11.73 (2.14) mCi, respectively, *W*=52, p=0.63.

Three striatal clusters showed ASD>control group differences for the contrast of (reward>neutral) BP_ND_ values, reflecting greater difference in phasic DA release in the reward relative to the neutral condition in the ASD group relative to control group. Clusters were located in the right putamen, left putamen, and left caudate nucleus/left putamen. Condition-specific BP_ND_ values were extracted from each participant. In all conditions, there was a significant Group × Condition interaction, *F’s*(1.20)> 8.6, *p*’s<.009 (see Figure 3 and Table 2). In general, the ASD control group exhibited increased BP_ND_ in the reward condition relative to the neutral condition, interpreted as decreased phasic DA release to rewards, whereas the control group showed the inverse response. Because the left caudate nucleus/left putamen cluster contained white matter voxels, this region was further analyzed and sub-regional analysis confirmed the presence of the effect in the grey matter of left caudate and putamen (described in **Supplemental Materials IV)**. Furthermore, region-of-interest analyses using the 1mm striatum structural atlas [55] showed a significant difference in the right putamen and trending differences in the left putamen and left caudate (presented in **Supplementary Materials V**), supporting the aforementioned SRTM results. Finally, of note, one cluster in the right caudate nucleus demonstrated the opposite pattern of group differences for the contrast of (reward>neutral) BP_ND_ values: relative to controls, the ASD group exhibited decreased BP_ND_ (or increased DA) in the reward relative to the neutral condition, Group × Condition interaction, *F*(1,20)= 7.5, *p*<.05.

**Figure 3.**
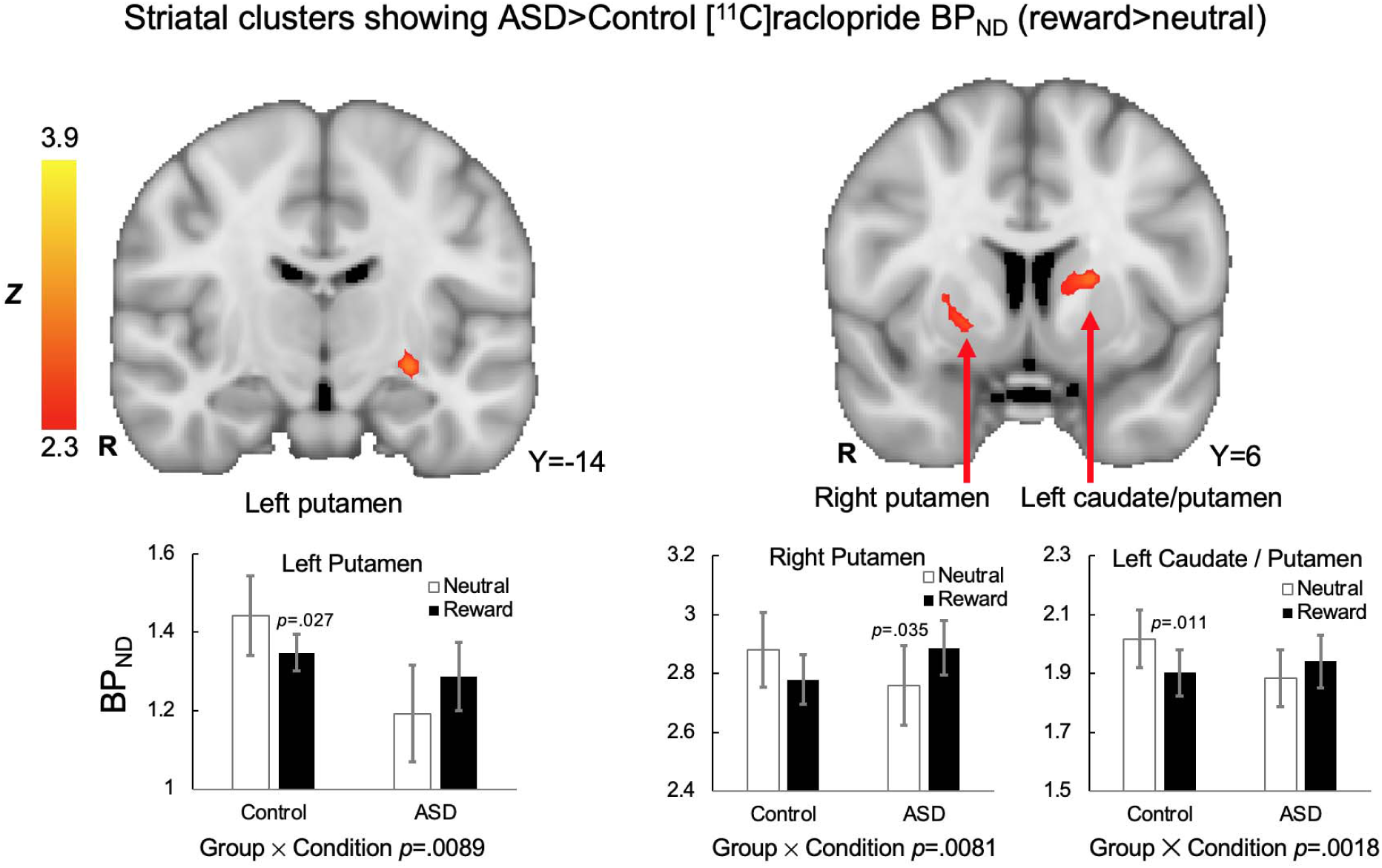
Striatal clusters that showed ASD>Control [^11^C]raclopride BP_ND_ (reward>neutral), signifying decreased phasic release of dopamine to rewards in the ASD group relative to the control group, were evident in the left putamen, right putamen, and a cluster that hat spanned the left caudate nucleus and putamen. For all clusters, the Group ⍰ Condition interaction effect on [^11^C]raclopride BP_ND_ values were significant. In the right putamen cluster, the ASD group demonstrated a pattern of greater phasic DA release to neutral than reward blocks.

**Table 2.** Striatal clusters demonstrated ASD>control group differences for the contrast of (reward>neutral) BP_ND_ values at the threshold of z>2.3, reflecting greater difference in phasic DA release in the reward relative to the neutral condition in the control group relative to the ASD group.

| **Cluster Label** | **Cluster Size**<br>**(voxels)** | **Max Z**<br>**value** | **Max X** | **Max Y** | **Max Z** |
| --- | --- | --- | --- | --- | --- |
| left caudate nucleus /<br>putamen | 87 | 3.95 | -18 | 2 | 10 |
| right putamen | 41 | 2.84 | 22 | 6 | -2 |
| left putamen | 40 | 3.22 | -26 | -16 | -8 |

### fMRI General Functional Connectivity (GFC) Results

Whole-brain GFC analysis revealed significant group differences in connectivity with the PET-derived right putamen seed (based on group differences for the contrast of [reward>neutral] BP_ND_ values), but no other PET-derived striatal seeds. Compared to the control group, the ASD group exhibited relatively greater connectivity between the right putamen seed and the precuneus and right insular cortex (see Figure 4 and Table 3).

**Figure 4.**
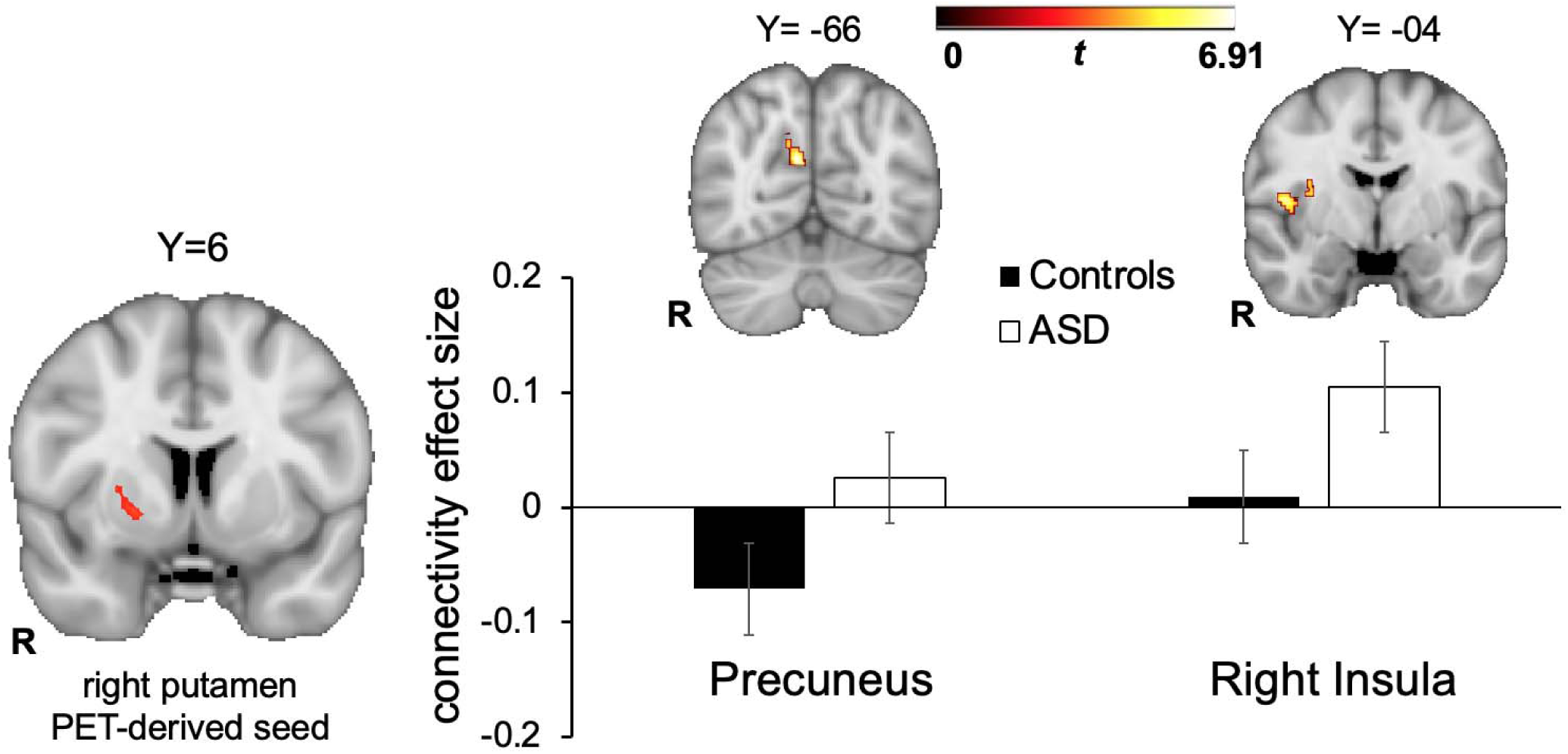
ASD>Control differences in general functional connectivity between the PET-derived right putamen seed and precuneus andnd right insula, displayed in MNI152 space at peak coordinates for each target region. The bar graph shows the effect size for each target region, represented by the Fisher-transformed correlation coefficients, separated by group.

**Table 3.**
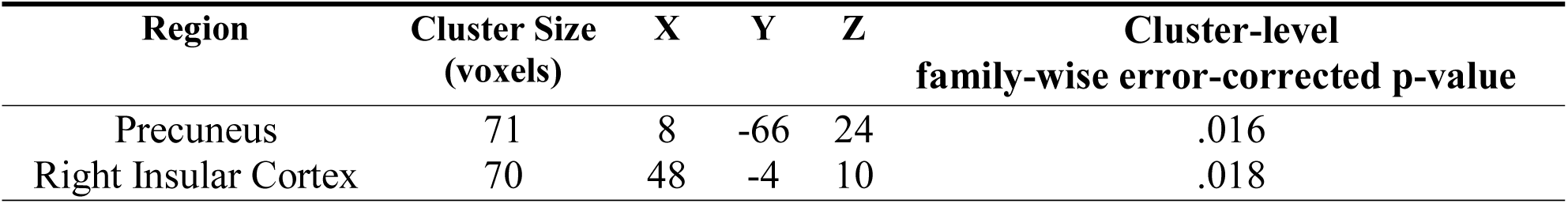
Regions showing greater connectivity in the ASD relative to the control group with the PET-derived right putamen seed that demonstrated group differences for (reward>neutral) BP_ND_ values.

| **Region** | **Cluster Size**<br>**(voxels)** | **X** | **Y** | **Z** | **Cluster-level**<br>**family-wise error-corrected p-value** |
| --- | --- | --- | --- | --- | --- |
| Precuneus | 71 | 8 | -66 | 24 | .016 |
| Right Insular Cortex | 70 | 48 | -4 | 10 | .018 |

### Supplementary fMRI Activation and Task-Based gPPI Results

fMRI activation results are described in **Supplemental Materials VI**. Results of secondary task-based gPPI analyses are presented in **Supplemental Materials VII**.

### Task Reaction Time and Valence Ratings

As shown in **Supplementary Materials VIII**, there were no significant group or group interaction effects for RT or valence ratings, *p’s* > 0.05, although these measures indicated the task elicited motivated behavior and positive subjective experience.

### Correlations between Striatal Dopamine Binding and ASD Symptom Severity

Exploratory correlational analyses in the ASD group considered associations between (reward>neutral) BP_ND_ values in the four striatal PET clusters that differentiated groups, and ADOS-2 calibrated severity scores [56], SRS total t-scores, and RMITE scores. Only one association emerged in the hypothesized direction: decreased phasic DA release to incentives in the left putamen was related to worse performance on the RMITE, a measure of theory-of-mind, in the ASD group (*p*=0.03; see Figure 5). This result was only significant at an uncorrected significance threshold.

**Figure 5.**
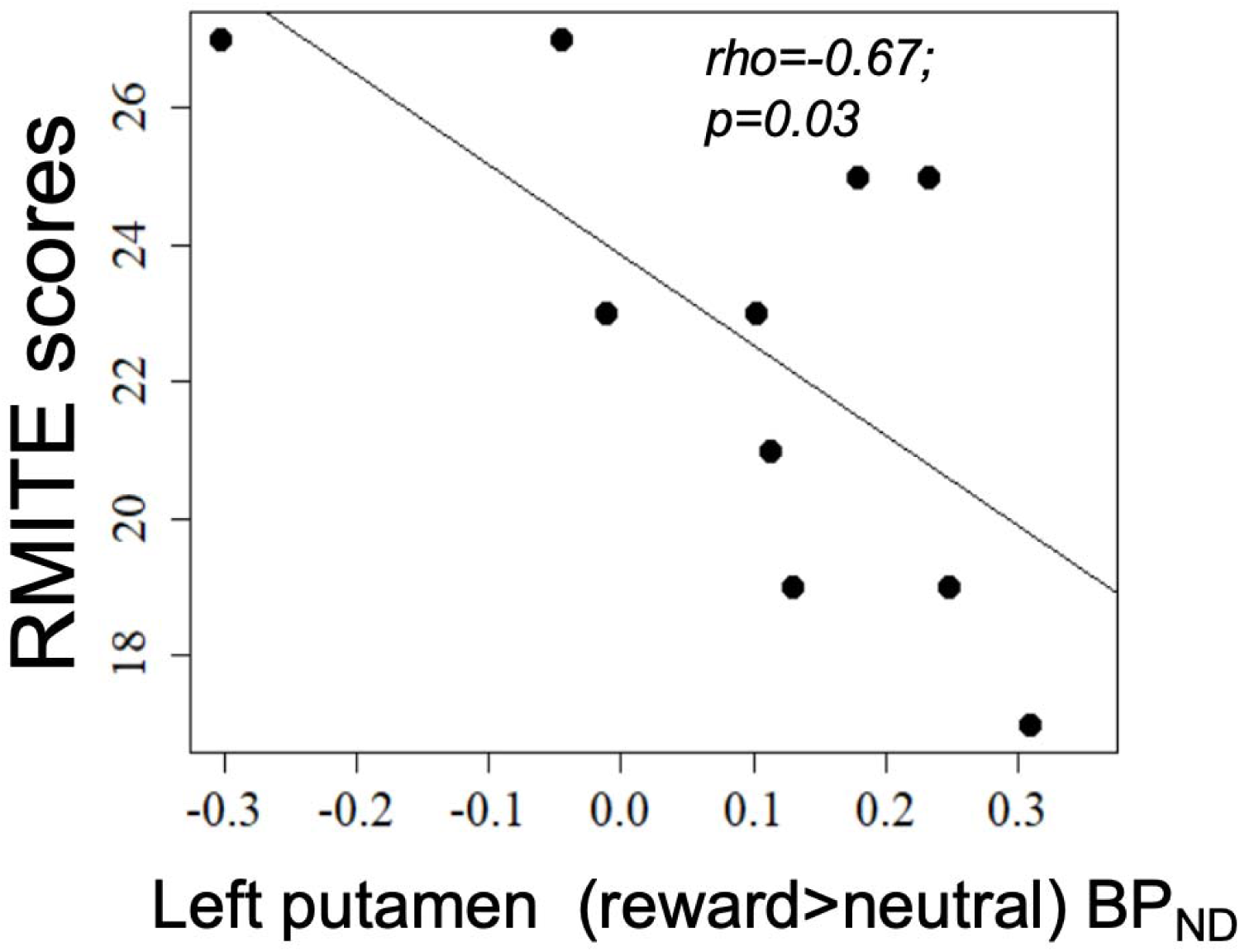
Association between left putamen (reward>neutral) BP_ND_ values and the Reading the Mind in the Eyes” Test, Revised Version (RMITE) scores in the ASD group. Higher RMITE scores indicate better performance. Higher BP_ND_ values indicate decreased phasic dopamine release to incentives.

## Discussion

The social motivation hypothesis of autism proposes that impaired reward circuitry responses to social information give rise to social communication symptoms in ASD. Although numerous fMRI, electrophysiological, and behavioral studies have investigated this framework [57], no previous molecular imaging study has directly investigated striatal DA functioning in ASD. In this study we evaluated striatal DA functioning in ASD via simultaneous PET and fMRI during incentive processing using the D2/D3 dopamine receptor antagonist [^11^C]raclopride and a novel MID task. We supplemented our PET analysis by using fMRI to examine functional connectivity of striatal regions that showed impaired DA release in ASD.

Analysis of [^11^C]raclopride PET data revealed relatively decreased phasic DA release to rewards in the ASD group in several striatal clusters, including the putamen and caudate nucleus. The putamen and the caudate nucleus comprise the dorsal striatum, a structure shown to be centrally involved in reinforcement learning and goal-directed behaviors that are facilitated by dorsal striatal DA release [58, 59]. Specifically, the dorsal striatum plays an important role in learning stimulus-action-outcome associations and stimulus-action coding [60]. Though the present study did not investigate reward learning, this pattern of impaired dorsal striatal DA release in the ASD group is consistent with the well-documented deficits in learning [61, 62] and flexible responses to environmental contingencies [63] in ASD that may result from atypical computation of prediction errors [64, 65]. The neuroimaging literature addressing reward learning in ASD has found decreased frontostriatal activity during both implicit and explicit social reward learning tasks [15, 66, 67], underscoring the potential relevance of impaired reward learning to core ASD symptoms.

fMRI functional connectivity with PET-derived striatal seed regions was evaluated with a general functional connectivity (GFC) approach. The only PET-derived seed region that showed group differences in GFC was the right putamen. Greater connectivity was observed in the ASD group between the right putamen and the precuneus and right insular cortex. The precuneus has direct connections to the basal ganglia [68], is involved in self-referential processing [69] and has been linked to mentalizing deficits in ASD [70]. Notably, increased striatal connectivity with the precuneus during reward processing has been associated with depressive symptom severity in anhedonic patients with major depressive disorder (MDD) [71]. Thus, in the present context, increased connectivity between a putamen cluster demonstrating decreased phasic DA release and the precuneus may reflect decreased motivation, indicating a possible shared feature of ASD and MDD.

The right putamen PET-derived seed also demonstrated increased connectivity with the right insular cortex in the ASD group. The insular cortex, and in particular its anterior portion, is a critical hub for regulating large-scale brain network dynamics [72] and is part of the salience network that integrates sensory, autonomic, and hedonic input to guide behavior [73]. In ASD, there is evidence that the insula plays a key role in social and non-social ASD impairments [74]. A meta-analysis of functional neuroimaging ASD studies found insula hypoactivation in ASD during a range of social processing tasks [75], suggesting that insula dysfunction may be central to the disorder [76] given the multiple functions subserved by the insula, including attention and affective processing of salient social information [77]. Increased functional connectivity between the putamen and insula has been reported during rest in children with ASD, though this pattern of abnormal insular connectivity was not specific to the putamen but rather observed in a number of dorsal and ventral striatal seeds and cortical regions [78]. The finding in the present context of increased connectivity between a putamen cluster demonstrating decreased phasic DA release to rewards and the right insular cortex highlights that striatal DA signals may drive the impaired functioning of various associative and limbic cortices implicated in the pathophysiology of ASD.

Exploratory fMRI activation in response to the incentive task revealed decreased activation in the left putamen during reward anticipation and in the anterior cingulate gyrus during reward outcomes in the ASD group, though the former finding was at an uncorrected threshold. Broadly, these results are consistent with the literature documenting decreased neural responses to monetary rewards in ASD using fMRI [12]. Exploratory gPPI connectivity analyses revealed ASD>control group differences during reward anticipation in connectivity between the PET-derived left putamen cluster that demonstrated group differences for the contrast of (reward>neutral) BP_ND_ values and the orbital frontal cortex, a region implicated in oxytocin response in ASD [26]. Finally, exploratory correlational analyses in the ASD group revealed that decreased phasic DA release to incentives in the left putamen was related to worse theory-of-mind, a core ASD impairment [79], highlighting the clinical significance of PET findings.

This study had several limitations. First, the sample sizes are small. Second, the imaging task presented monetary rewards rather than social rewards. This design was to ensure a robust striatal DA response in the control group given the extant literature demonstrating striatal DA release to monetary rewards in nonclinical samples using [^11^C]raclopride [e.g., 80, 81]. Although the social motivation hypothesis of autism highlights impaired responses to social rewards in ASD, several studies report striatal dysfunction to both social and non-social rewards in ASD [12, 57, 82, 83]. Thus, the present study has mechanistic relevance to address impaired social motivation responses in ASD. This study was also restricted to participants with higher cognitive abilities and may not represent the broader ASD population. In this regard, we have established PET-MR protocols for adults with ASD with lower intellectual functioning [84] and recently completed a PET-MR study that included individuals with ASD with full-scale IQs ranging from 47 to 112 [85]. Finally, two results should be interpreted with caution until replicated: 1) a cluster in the right caudate nucleus showed increased phasic DA release to rewards in the ASD group, a finding that was unexpected and in the opposite direction of other striatal PET clusters; and 2) the relation between phasic DA release in the left putamen and performance on the theory-of-mind measure was only significant at an uncorrected threshold.

In spite of these limitations, this study is the first PET-MR investigation of striatal DA functioning in ASD. Using [^11^C]raclopride in conjunction with a reward processing task, we report evidence consistent with impaired phasic DA release to rewards in the striatum in ASD. We further demonstrated that functional connectivity in the ASD group was increased between a PET-derived right putamen seed (that exhibited decreased phasic DA release to rewards) and the precuneus and right insula, suggesting a molecular mechanism that may address, in part, the pathogenisis of impaired functional brain networks in ASD. These results indicate that ASD is characterized by impaired striatal DA functioning, consistent with the social motivation hypothesis of autism, and highlights that PET-MR may be a suitable tool to evaluate novel treatments aimed at improving striatal DA functioning in ASD. More broadly, the use of simultaneous PET-MR represents an important means to address the heterogeneity of ASD [86] by identifying individuals characterized by homogenous molecular etiologies. It additionally holds the promise of validating the molecular underpinnings of fMRI signals. Finally, it provides perhaps the most direct linkages possible between human disorders and preclinical animal models characterized by common molecular pathophysiologies [87].

## Data Availability

The data from this study are available for download from the National Database for Autism Research (NDAR), collection #2471

## Acknowledgements

This research was supported by R21 MH110933 to GSD and JMH, K23 MH113733 to ECW, and UL1TR002489. Assistance with recruitment was provided by the Clinical Translational Core of the UNC Intellectual Developmental Disabilities Research Center (HD103573). The data from this study are available for download from the National Database for Autism Research (NDAR), collection #2471. We would like to thank Grae Arabasz for technical advice for PET-MR acquisition. We extend our sincere gratitude to the individuals who participated in this study.

## References

1. Chevallier, C., et al., The Social Motivation Theory of Autism. Trends in Cognitive Sciences, 2012. 16(4): p. 231–239.

2. Dawson, G., S.J. Webb, and J. McPartland, Understanding the nature of face processing impairment in autism: insights from behavioral and electrophysiological studies. Dev Neuropsychol, 2005. 27(3): p. 403–24.

3. Kuhl, P.K., et al., Links between social and linguistic processing of speech in preschool children with autism: behavioral and electrophysiological measures. Dev Sci, 2005. 8(1): p. F1–F12.

4. Berridge, K.C. and T.E. Robinson, Parsing reward. Trends Neurosci, 2003. 26(9): p. 507–13.

5. Berridge, K.C., T.E. Robinson, and J.W. Aldridge, Dissecting components of reward: ‘liking’, ‘wanting’, and learning. Curr Opin Pharmacol, 2009. 9(1): p. 65–73.

6. Schultz, W., Predictive reward signal of dopamine neurons. J Neurophysiol, 1998. 80(1): p. 1–27.

7. Gunaydin, L.A. and K. Deisseroth, Dopaminergic Dynamics Contributing to Social Behavior. Cold Spring Harb Symp Quant Biol, 2014. 79: p. 221–7.

8. Gu, R., et al., Love is analogous to money in human brain: Coordinate-based and functional connectivity meta-analyses of social and monetary reward anticipation. Neurosci Biobehav Rev, 2019. 100: p. 108–128.

9. Manduca, A., et al., Dopaminergic Neurotransmission in the Nucleus Accumbens Modulates Social Play Behavior in Rats. Neuropsychopharmacology, 2016.

10. Dichter, G.S., et al., Reward circuitry function in autism during face anticipation and outcomes. J Autism Dev Disord, 2012. 42(2): p. 147–60.

11. Mosner, M.G., et al., Neural Mechanisms of Reward Prediction Error in Autism Spectrum Disorder. Autism Res Treat, 2019. 2019: p. 5469191.

12. Clements, C.C., et al., Evaluation of the Social Motivation Hypothesis of Autism: A Systematic Review and Meta-analysis. JAMA Psychiatry, 2018. 75(8): p. 797–808.

13. Dichter, G.S., Motivational Impairments in Autism May Be Broader Than Previously Thought. JAMA Psychiatry, 2018. 75(8): p. 773–774.

14. Abrams, D.A., L.Q. Uddin, and V. Menon, Reply to Brock: Renewed focus on the voice and social reward in children with autism. Proc Natl Acad Sci U S A, 2013. 110(42): p. E3974.

15. Choi, U.S., et al., Abnormal brain activity in social reward learning in children with autism spectrum disorder: an fMRI study. Yonsei Med J, 2015. 56(3): p. 705–11.

16. Dichter, G.S., C.A. Damiano, and J.A. Allen, Reward circuitry dysfunction in psychiatric and neurodevelopmental disorders and genetic syndromes: animal models and clinical findings. J Neurodev Disord, 2012. 4(1): p. 19.

17. Marotta, R., et al., The Neurochemistry of Autism. Brain Sci, 2020. 10(3).

18. Mosner, M.G., et al., Vicarious Effort-Based Decision-Making in Autism Spectrum Disorders. J Autism Dev Disord, 2017. 47(10): p. 2992–3006.

19. Gadow, K.D., et al., Association of DRD4 polymorphism with severity of oppositional defiant disorder, separation anxiety disorder and repetitive behaviors in children with autism spectrum disorder. Eur J Neurosci, 2010. 32(6): p. 1058–65.

20. Gadow, K.D., et al., Parent-child DRD4 genotype as a potential biomarker for oppositional, anxiety, and repetitive behaviors in children with autism spectrum disorder. Prog Neuropsychopharmacol Biol Psychiatry, 2010. 34(7): p. 1208–14.

21. Staal, W.G., Autism, DRD3 and repetitive and stereotyped behavior, an overview of the current knowledge. Eur Neuropsychopharmacol, 2015. 25(9): p. 1421–6.

22. Dolen, G., Autism: Oxytocin, serotonin, and social reward. Soc Neurosci, 2015. 10(5): p. 450–65.

23. Depue, R. and J. Morrone-Strupinsky, A neurobehavioral model of affiliative bonding: implications for conceptualizing a human trait of affiliation. The Behavioral and brain sciences, 2005. 28(3): p. 313.

24. Parker, K.J., et al., Intranasal oxytocin treatment for social deficits and biomarkers of response in children with autism. Proc Natl Acad Sci U S A, 2017. 114(30): p. 8119–8124.

25. Higashida, H., et al., Social Interaction Improved by Oxytocin in the Subclass of Autism with Comorbid Intellectual Disabilities. Diseases, 2019. 7(1).

26. Greene, R.K., et al., The effects of intranasal oxytocin on reward circuitry responses in children with autism spectrum disorder. J Neurodev Disord, 2018. 10(1): p. 12.

27. Ferguson, J.N., L.J. Young, and T.R. Insel, The neuroendocrine basis of social recognition. Frontiers in neuroendocrinology, 2002. 23(2): p. 200–224.

28. Olazabal, D. and L. Young, Oxytocin receptors in the nucleus accumbens facilitate “spontaneous” maternal behavior in adult female prairie voles. Neuroscience, 2006. 141(2): p. 559–568.

29. Dolen, G., et al., Social reward requires coordinated activity of nucleus accumbens oxytocin and serotonin. Nature, 2013. 501(7466): p. 179–84.

30. Choe, H.K., et al., Oxytocin Mediates Entrainment of Sensory Stimuli to Social Cues of Opposing Valence. Neuron, 2015. 87(1): p. 152–63.

31. Mabunga, D.F., et al., Exploring the Validity of Valproic Acid Animal Model of Autism. Exp Neurobiol, 2015. 24(4): p. 285–300.

32. Kuo, H.Y. and F.C. Liu, Molecular Pathology and Pharmacological Treatment of Autism Spectrum Disorder-Like Phenotypes Using Rodent Models. Front Cell Neurosci, 2018. 12: p. 422.

33. Narita, N., et al., Increased monoamine concentration in the brain and blood of fetal thalidomide- and valproic acid-exposed rat: putative animal models for autism. Pediatr Res, 2002. 52(4): p. 576–9.

34. Nakasato, A., et al., Swim stress exaggerates the hyperactive mesocortical dopamine system in a rodent model of autism. Brain Res, 2008. 1193: p. 128–35.

35. Shaywitz, B.A., R.D. Yager, and J.H. Klopper, Selective brain dopamine depletion in developing rats: an experimental model of minimal brain dysfunction. Science, 1976. 191(4224): p. 305–8.

36. First, M.B., et al., Structured Clinical Interview for DSM-5-Research Version (SCID-5 for DSM-5, Research Version; SCID-5-RV). 2015, American Psychiatric Association: Arlington, VA.

37. Chandler, S., et al., Validation of the social communication questionnaire in a population cohort of children with autism spectrum disorders. J Am Acad Child Adolesc Psychiatry, 2007. 46(10): p. 1324–32.

38. Uttl, B., North American Adult Reading Test: age norms, reliability, and validity. J Clin Exp Neuropsychol, 2002. 24(8): p. 1123–37.

39. Axelrod, B.N., Validity of the Wechsler abbreviated scale of intelligence and other very short forms of estimating intellectual functioning. Assessment, 2002. 9(1): p. 17–23.

40. Baron-Cohen, S., et al., The "Reading the Mind in the Eyes" Test revised version: a study with normal adults, and adults with Asperger syndrome or high-functioning autism. J Child Psychol Psychiatry, 2001. 42(2): p. 241–51.

41. Hus, V., K. Gotham, and C. Lord, Standardizing ADOS domain scores: separating severity of social affect and restricted and repetitive behaviors. J Autism Dev Disord, 2014. 44(10): p. 2400–12.

42. Watabe, H., et al., Measurement of dopamine release with continuous infusion of [11C]raclopride: optimization and signal-to-noise considerations. J Nucl Med, 2000. 41(3): p. 522–30.

43. Izquierdo-Garcia, D., et al., An SPM8-based approach for attenuation correction combining segmentation and nonrigid template formation: application to simultaneous PET/MR brain imaging. J Nucl Med, 2014. 55(11): p. 1825–30.

44. Knutson, B., et al., FMRI visualization of brain activity during a monetary incentive delay task. Neuroimage, 2000. 12(1): p. 20–7.

45. Peirce, J., et al., PsychoPy2: Experiments in behavior made easy. Behav Res Methods, 2019. 51(1): p. 195–203.

46. Knutson, B., et al., A region of mesial prefrontal cortex tracks monetarily rewarding outcomes: characterization with rapid event-related fMRI. Neuroimage, 2003. 18(2): p. 263-72

47. Sander, C.Y., et al., Imaging Agonist-Induced D2/D3 Receptor Desensitization and Internalization in vivo with PET/fMRI. Neuropsychopharmacology, 2015.

48. Sander, C.Y., et al., Imaging Agonist-Induced D2/D3 Receptor Desensitization and Internalization In Vivo with PET/fMRI. Neuropsychopharmacology, 2016. 41(5): p. 1427–36.

49. Elliott, M.L., et al., General functional connectivity: Shared features of resting-state and task fMRI drive reliable and heritable individual differences in functional brain networks. 263-72. Neuroimage, 2019. 189: p. 516–532.

50. Anderson, J.S., et al., Reproducibility of single-subject functional connectivity measurements. AJNR Am J Neuroradiol, 2011. 32(3): p. 548–55.

51. Hacker, C.D., et al., Resting state network estimation in individual subjects. Neuroimage, 2013. 82: p. 616–633.

52. Laumann, T.O., et al., Functional System and Areal Organization of a Highly Sampled Individual Human Brain. Neuron, 2015. 87(3): p. 657–70.

53. Whitfield-Gabrieli, S. and A. Nieto-Castanon, Conn: a functional connectivity toolbox for correlated and anticorrelated brain networks. Brain Connect, 2012. 2(3): p. 125–41.

54. Nieto-Castanon, A., Handbook of functional connectivity Magnetic Resonance Imaging methods in CONN. 2020, Boston, MA: Hilbert Press.

55. Tziortzi, A.C., et al., Imaging dopamine receptors in humans with [11C]-(+)-PHNO: dissection of D3 signal and anatomy. Neuroimage, 2011. 54(1): p. 264–77.

56. Gotham, K., A. Pickles, and C. Lord, Standardizing ADOS scores for a measure of severity in autism spectrum disorders. J Autism Dev Disord, 2009. 39(5): p. 693–705.

57. Bottini, S., Social reward processing in individuals with autism spectrum disorder: a systematic review of the social motivation hypothesis. Research in Autism Spectrum Disorders, 2018. 45: p. 9–26.

58. Packard, M.G. and B.J. Knowlton, Learning and memory functions of the Basal Ganglia. Annu Rev Neurosci, 2002. 25: p. 563–93.

59. Graybiel, A.M., Building action repertoires: memory and learning functions of the basal ganglia. Curr Opin Neurobiol, 1995. 5(6): p. 733–41.

60. O’Doherty, J., et al., Dissociable roles of ventral and dorsal striatum in instrumental conditioning. Science, 2004. 304(5669): p. 452–4.

61. Lin, A., A. Rangel, and R. Adolphs, Impaired learning of social compared to monetary rewards in autism. Front Neurosci, 2012. 6: p. 143.

62. Mussey, J.L., et al., Decision-making skills in ASD: performance on the Iowa Gambling Task. Autism Res, 2015. 8(1): p. 105–14.

63. Sinha, P., et al., Autism as a disorder of prediction. Proc Natl Acad Sci U S A, 2014. 111(42): p. 15220–5.

64. Lawson, R.P., G. Rees, and K.J. Friston, An aberrant precision account of autism. Front Hum Neurosci, 2014. 8: p. 302.

65. Van de Cruys, S., et al., Precise minds in uncertain worlds: predictive coding in autism. Psychol Rev, 2014. 121(4): p. 649–75.

66. Scott-Van Zeeland, A.A., et al., Reward processing in autism. Autism Res, 2010. 3(2): p. 53–67.

67. Kinard, J.L., et al., Neural Mechanisms of Social and Nonsocial Reward Prediction Errors in Adolescents with Autism Spectrum Disorder. Autism Res, 2020.

68. Cavanna, A.E., The precuneus and consciousness. CNS Spectr, 2007. 12(7): p. 545–52.

69. Dammann, G., et al., The self-image in borderline personality disorder: an in-depth qualitative research study. J Pers Disord, 2011. 25(4): p. 517–27.

70. Mundy, P., A review of joint attention and social-cognitive brain systems in typical development and autism spectrum disorder. Eur J Neurosci, 2018. 47(6): p. 497–514.

71. Quevedo, K., et al., Ventral Striatum Functional Connectivity during Rewards and Losses and Symptomatology in Depressed Patients. Biol Psychol, 2017. 123: p. 62–73.

72. Uddin, L.Q., Salience processing and insular cortical function and dysfunction. Nat Rev Neurosci, 2015. 16(1): p. 55–61.

73. Seeley, W.W., et al., Dissociable intrinsic connectivity networks for salience processing and executive control. J Neurosci, 2007. 27(9): p. 2349–56.

74. Nomi, J.S., I. Molnar-Szakacs, and L.Q. Uddin, Insular function in autism: Update and future directions in neuroimaging and interventions. Prog Neuropsychopharmacol Biol Psychiatry, 2019. 89: p. 412–426.

75. Di Martino, A., et al., Relationship between cingulo-insular functional connectivity and autistic traits in neurotypical adults. Am J Psychiatry, 2009. 166(8): p. 891–9.

76. Uddin, L.Q. and V. Menon, The anterior insula in autism: under-connected and under-examined. Neurosci Biobehav Rev, 2009. 33(8): p. 1198–203.

77. Odriozola, P., et al., Insula response and connectivity during social and non-social attention in children with autism. Soc Cogn Affect Neurosci, 2016. 11(3): p. 433–44.

78. Di Martino, A., et al., Aberrant striatal functional connectivity in children with autism. Biol Psychiatry, 2011. 69(9): p. 847–56.

79. Association”, A.P., Diagnostic and statistical manual of mental disorders. 5th ed. 2013, Arlington, VA: American Psychiatric Publishing.

80. Zald, D.H., et al., Dopamine transmission in the human striatum during monetary reward tasks. J Neurosci, 2004. 24(17): p. 4105–12.

81. Pappata, S., et al., In vivo detection of striatal dopamine release during reward: a PET study with [(11)C]raclopride and a single dynamic scan approach. Neuroimage, 2002. 16(4): p. 1015–27.

82. Dichter, G.S., et al., Reward circuitry function in autism spectrum disorders. Soc Cogn Affect Neurosci, 2012. 7(2): p. 160–72.

83. Cascio, C.J., et al., Affective neural response to restricted interests in autism spectrum disorders. J Child Psychol Psychiatry, 2014. 55(2): p. 162–71.

84. Smith, C.J., et al., A Protocol for Sedation Free MRI and PET Imaging in Adults with Autism Spectrum Disorder. J Autism Dev Disord, 2019. 49(7): p. 3036–3044.

85. Zurcher, N.R., et al., [(11)C]PBR28 MR-PET imaging reveals lower regional brain expression of translocator protein (TSPO) in young adult males with autism spectrum disorder. Mol Psychiatry, 2020.

86. Masi, A., et al., An Overview of Autism Spectrum Disorder, Heterogeneity and Treatment Options. Neurosci Bull, 2017. 33(2): p. 183–193.

87. van der Doef, T.F., et al., Assessing brain immune activation in psychiatric disorders: clinical and preclinical PET imaging studies of the 18-kDa translocator protein. Clin Transl Imaging, 2015. 3(6): p. 449–460.

88. Constantino, J.N. and C.P. Gruber, The social responsiveness scale. Los Angeles: Western Psychological Services. 2002.

89. Hollingshead, A., Four factor index of social status. 1975, Yale University Department of Psychology: New Haven.

